# Understanding Mental Health in Crisis: Key Determinants of Psychological Distress in Belgium during the first weeks of COVID-19 Lockdown

**DOI:** 10.64898/2026.07.15.26358143

**Authors:** Z. Zsabokorszky, K. Pepermans, K. Van den Broeck, P. Beutels, N. Hens

## Abstract

**Aims:** The COVID-19 pandemic has significantly impacted global mental health. At the onset of the pandemic (2020), Belgians experienced increased anxiety, depression, and psychological distress compared to 2018 due to the outbreak and the associated public health measures. Understanding the drivers of this distress is crucial for mitigating mental health effects in future crises. This study examines determinants of psychological distress in Belgium during the March 2020 lockdown, using data from the Great Corona Study (GCS).

**Methods:** Data were drawn from the second wave of the GCS, a citizen science initiative conducted in Belgium on March 24, 2020, with 332,169 respondents. Psychological distress was measured using the General Health Questionnaire-12 (GHQ-12), applying a 2/3 cutoff to classify distress levels. To identify predictor variables, a random forest algorithm and literature review reduced 207 initial variables to 16. A generalized linear model was then used to examine associations between predictors and psychological distress

**Results:** Psychological distress was significantly associated with various demographic, social, occupational, and health-related factors. Younger individuals, women, and residents of Wallonia or Brussels exhibited higher odds of distress. Household composition, and the frequency of real-life social interactions significantly influenced distress levels. Occupational status played a key role, with part-time employees and working students exhibiting higher levels of distress. At the same time retired individuals with no current occupation showed lower odds. Perceived workplace safety and compliance with public health measures also significantly impacted distress levels. Lastly, individuals experiencing influenza-like or COVID-19 symptoms had substantially higher odds of psychological distress.

**Conclusions:** Our findings highlight significant sociodemographic, occupational, and health-related predictors of psychological distress during the initial COVID-19 lockdown in Belgium. Young adults, women, individuals with limited in-person interactions, and those experiencing influenza-like illness or COVID-19 symptoms were particularly vulnerable. Additionally, perceptions of others’ adherence to preventive measures played a crucial role in mental well-being. These results highlight the complex interplay between individual and environmental factors in shaping psychological distress, providing valuable insights for future public health policies and mental health interventions during crises.

## Background

The COVID-19 pandemic has had a profound impact on mental health worldwide (1, 2). According to the World Health Organization, global prevalence of depression and anxiety increased by 25% in the first year of the pandemic due to pandemic-related stressors (2). In Belgium, this global trend was also evident during the early stages of the pandemic in 2020, as the outbreak and the corresponding public health measures led to a significant rise in anxiety, depression, and psychological distress (3–9).

During the initial lockdown phase, which this study focuses on, pressure on the Belgian hospital system was comparable to other European countries, though notably lower than in Italy, Spain, and the Netherlands (10). The first confirmed COVID-19 case in Belgium was reported on February 3, 2020, with the first death occurring on March 10. In response, the government implemented emergency measures, including school, business, and border closures, by enforcing a nationwide lockdown by mid-March. While important to contain viral transmission, these restrictions disrupted daily life, caused social isolation, and resulted in economic hardship (11).

Several sociodemographic factors have since then been linked to increased risk of anxiety and depression, including education level, professional situation and its disruption due to the pandemic, and household composition (4). Moreover, direct or indirect contact with infected or suspected cases increased the risk of developing anxiety or depressive disorders (4). Similarly, contracting COVID-19 and experiencing its symptoms were associated with poorer mental health outcomes (3).

Certain personality traits, like extraversion, along with pre-existing psychological conditions further increased vulnerability (3, 6). Additionally, loneliness, increased social media use, and decreased in-person social interactions worsened mental distress during this period (9). Conversely, support from loved ones and the social environment played a protective role (3, 4, 8, 9). Finally, the degree to which various aspects of life were disrupted by the pandemic was strongly linked to worsening mental health (4).

Certain occupational groups were especially vulnerable, with healthcare workers experiencing exceptional psychological distress during the early lockdown (March 2020) (12).The mental health impact was also particularly pronounced among children, adolescents, and students, who experienced significant psychological distress during the pandemic, while young adults showed increased mental health care needs during the first pandemic year (9, 13). The elderly population faced substantial challenges as well, especially those in institutional settings. Higher levels of depression were associated with declines in daily activities, reduced sleep quality and well-being, and cognitive dysfunction (14).

Extensive research has identified various factors associated with mental health outcomes, providing valuable insights into psychological distress during crises. This study leverages data from the Great Corona Study (GCS) to examine these associations in the unique context of the initial COVID-19 lockdown in Belgium.

The GCS, a citizen science project, aimed to gather real-time data on how the Belgian population was coping with the COVID-19 crisis. The recurrent non-probabilistic, observational survey was conducted weekly between March 10, 2020, and March 29, 2022, with sample sizes ranging from 13,000 to 560,000 per wave. Launched by scientists at the University of Antwerp in collaboration with Hasselt University and KU Leuven, the study provided critical population-level insights in such aspects of the pandemic as the spread of COVID-19 symptoms and the economic and societal impact of the pandemic, and addressed an urgent need for indicative data to monitor well-being of the general population, i.e. not only subgroups of the general population (15).

While previous articles utilizing GCS data primarily focused on other aspects of the pandemic, like the spatial distribution and prediction of future infections (16), adaptations in consumer behavior (17), attitudes towards vaccine acceptance (18), drivers of compliance with COVID-19 measures (19), and overall health and wellbeing impacts (20), the psychological dimension has not yet been fully explored. This study aims to address this gap by examining predictors of psychological distress using GCS data from the initial COVID-19 lockdown in Belgium. The GCS dataset offers several advantages: its unprecedented scale, with a vast number of respondents; its exceptionally broad range of potential predictor variables; and its real-time and longitudinal data collection, capturing population dynamics as the crisis unfolded. By analyzing this dataset, we aim to gain new insights into the early mental health impact of lockdown measures.

## Methods

We used data from the second wave of the GCS, which was conducted on 24th March 2020 and retrieved information from 332,169 respondents.

Besides sociodemographic and economic variables, Covid-19 symptoms, indicators of physical and mental well-being, the questionnaire included the General Health Questionnaire-12 (GHQ-12) as a measurement of psychological distress. This aggregated questionnaire score was used with a cutoff score of 2/3 to distinguish between high and low levels of distress. The GHQ-12 is a widely used short-form screening instrument designed to assess psychological distress in various populations. The GHQ-12 has been extensively validated as a robust screening tool for psychological distress, demonstrating high sensitivity and specificity across diverse populations and settings. Its validity has been confirmed in multiple languages and cultural contexts, with an average area under the ROC curve of 0.88, indicating strong discriminatory power. Higher GHQ-12 scores correspond to greater levels of psychological distress, reinforcing its effectiveness as a brief yet reliable measure in mental health research (21). It has been shown to produce results comparable to longer versions of the GHQ. The GHQ-12 has demonstrated strong validity in multiple studies, including a WHO investigation comparing its performance to the GHQ-28 (21).

To identify the most relevant predictor variables for model selection to the dataset containing 207 variables, we initially considered factor analysis; however, given that the majority of our variables were discrete, this approach was considered less appropriate. Instead, we employed the random forest methodology to assess variable importance in relation to the outcome variable, allowing for a data-driven selection of key predictors. Figure 1-4 in Appendix A present the Variable Importance Plots corresponding to the Random Forest Analysis conducted with 1.000 and 10.000 random observations. In addition, we also reviewed existing literature to ensure that theoretically relevant variables were considered. This combined strategy facilitated the reduction of dimensionality while preserving the most informative predictors for subsequent statistical analyses.

To determine the association between the selected predictor variables and the outcome, we employed a generalized linear model using the gamlss package in R (22). For model selection, we utilized the stepGAIC function, which optimizes model fit based on the Generalized Akaike Information Criterion (GAIC). Given the characteristics of the dataset, population weights were constructed to enhance the representativeness of the sample. These weights were derived from population distributions for age, gender, location (province), and educational level. By applying these adjustments, we aimed to mitigate potential biases arising from over- or underrepresentation of certain demographic groups, ensuring that the findings more accurately reflect the broader population.

Data analysis was conducted using R version 4.3.2 (22).

## Results

### Characteristics of the study sample

The original dataset contained 345,966 observations. After excluding missing values, a total of 332,169 observations were included in the statistical analysis using the gamlss package. From the initial set of 16 predictor variables identified through random forest analysis and literature review, the selection process resulted in a final model comprising 11 variables (*see Appendix A for the Variable Importance Plot corresponding to the random forest analysis*).The mean GHQ-12 score was 3.657 (SD = 3.187). Among the 332,169 observations, 150,817 (45.4%) were categorized with a “low” GHQ-12 score, based on the 2/3 cutoff points, and 181,352 (54.6%) obtained a “high” GHQ-12 score. Table 1 in Appendix B presents the distribution of the binary GHQ-12 scores across the predictor variables included in the analysis.

### Results of the generalized linear model

#### Demographic characteristics

Various demographic characteristics were significantly associated with different levels of psychological distress, measured by the GHQ-12 questionnaire. Younger participants were more likely to be at risk of experiencing psychological distress, with each additional year of age associated with a 1% significant decrease in the odds of high distress (OR = 0.989, 95% CI (0.988 - 0.989)).

Gender also played a significant role, as women had 63% higher odds of experiencing psychological distress compared to men (OR = 1.625, 95% CI (1.593 - 1.657)).

Regional differences were prominent, with residence in Wallonia or Brussels associated with significantly increased odds of distress compared to Flanders. Living in Wallonia was linked to an 82% increase in odds (OR =1.815, 95% CI (1.766 - 1.866)), while residing in Brussels was associated with 74% higher odds of experiencing psychological distress (OR = 1.740, 95% CI (1.678 - 1.804)).

At the same time, educational background was only associated with psychological distress at certain levels. Individuals with a professional bachelor’s degree had 5% higher odds (OR = 1.053, 95% CI (1.008 - 1.101)), while individuals with an academic bachelor’s or master’s degree had 18% higher odds (OR = 1.178, 95% CI (1.125-1.234)) of experiencing high psychological distress. Those with a postgraduate qualification showed a 24% increase in odds (OR = 1.244, 95% CI (1.148 - 1.347)) compared to those with primary education. No significant association was observed for individuals with a secondary education.

#### Social and household composition

Household composition, particularly the number of cohabitants, emerged as a significant factor influencing the likelihood of experiencing high psychological distress. Individuals living with two cohabitants had 8% higher odds of experiencing psychological distress compared to those living with one (OR = 1.077, 95% CI (1.044-1.112)). Living with 11–20 cohabitants was associated with a 30% increase (OR = 1.297, 95% CI (1.008 - 1.668)), while living alone with a 25% increase in odds (OR = 1.250, 95% CI (1.215 - 1.287)). No significant associations were observed for individuals with three or four cohabitants, with 5–10 cohabitants, or with more than 20 cohabitants.

Similarly, the presence of children in the household also played an important role. Participants living with two children exhibited 17% higher odds of psychological distress compared to those with one child (OR = 1.165, 95% CI (1.121 - 1.211)). Living with three children was associated with a 17% increase in odds (OR = 1.165, 95% CI (1.095 - 1.239)), while living with 4 was associated with 13% (OR = 1.134, 95% CI (1.004 - 1.282)). At the same time, individuals without children had 6% lower odds of experiencing psychological distress compared to those living with one child (OR = 0.944, 95% CI (0.915 - 0.974)). No significant associations were identified for individuals living with more than four children.

Furthermore, the number of real life (not by phone, face time, Skype, etc) conversation partners other than household members was significantly and inversely associated with psychological distress. Participants who interacted with two people had 8% lower odds of experiencing psychological distress (OR = 0.923, 95% CI (0.892 - 0.955)) compared to those participants who only interacted with one. Similarly, interacting with three people was associated with a 7% reduction (OR = 0.930, 95% CI (0.892 - 0.970)), four people with an 8% reduction (OR = 0.922, 95% CI (0.876 - 0.970)), five to ten people with an 8% reduction (OR =0.917, 95% CI (0.881 - 0.955)), and 11 to 20 people with a 6% reduction (OR = 0.936, 95% CI (0.884 - 0.992)). No significant associations were observed for individuals living alone.

#### Occupation and compliance with regulations

Activity and employment status were significantly associated with psychological distress during the study period. Part-time employees had 4% higher odds of experiencing high psychological distress compared to full-time employees (OR = 1.038, 95% CI (1.006 – 1.072)).

Participants with a mixed occupational background, whether currently unemployed, working part-time, or working full-time, showed no statistically significant association with psychological distress levels.

Retired participants showed a notable negative association with psychological distress. Retired individuals with no current employment had 26% lower odds of experiencing high psychological distress compared to full-time employees (OR = 0.737, 95% CI (0.707 – 0.768)), while those working part-time showed no significant association.

In contrast, student status was associated with elevated distress levels, particularly when combined with employment. Students without current employment showed no significant difference from full-time workers, whereas students working part-time had 44% higher odds of psychological distress (OR = 1.435, 95% CI (1.238 – 1.663)), and those working full-time had 26% higher odds (OR = 1.258, 95% CI (1.102 – 1.436)).

The location of work and perceived safety of the work environment were significant determinants of psychological distress. Compared to individuals who did not work on the 20th of March (reference category), participants working outside the home but with limited contact with others had 15% lower odds of experiencing high psychological distress (OR =0.854, 95% CI (0.801 - 0.910)). In contrast, those working outside of the home in close proximity to colleagues perceived as not taking adequate precautions had 40% higher odds of high distress (OR = 1.392, 95% CI (1.301 - 1.490)). Interestingly, participants who worked outside the home but perceived their coworkers as adequately cautious experienced a 17% reduction in odds of high distress (OR = 0.834, 95% CI (0.801 - 0.869)). Those working from home also showcased significantly lower odds of high psychological distress compared to non-workers, with a 4% reduction (OR =0.964, 95% CI (0.938 - 0.991)).

Finally, perceptions of compliance with public health recommendations at public places were also associated significantly with psychological distress. For each one-point increase on a 1–10 scale estimating others’ behavioral adaptation to the recommendations, participants showed a 5% decrease in the odds of experiencing high psychological distress (OR = 0.954, 95% CI (0.949 - 0.959)).

#### Influenza-Like Illness (ILI) and COVID-19 Symptoms

The presence of diagnostic symptoms consistent with Influenza-Like Illness (ILI) and COVID-19 were strongly associated with psychological distress. Individuals reporting ILI symptoms had 2.2 times higher odds of experiencing high psychological distress compared to those without symptoms (OR = 2.244, 95% CI (1.978 - 2.546)). Similarly, participants displaying characteristic symptoms of COVID-19 had 2.1 times higher odds of experiencing psychological distress compared to those without these symptoms (OR =2.072, 95% CI (1.742 - 2.464)).

Table 2 presents the results of the generalized linear model applying survey weights.

**Table 2-.**
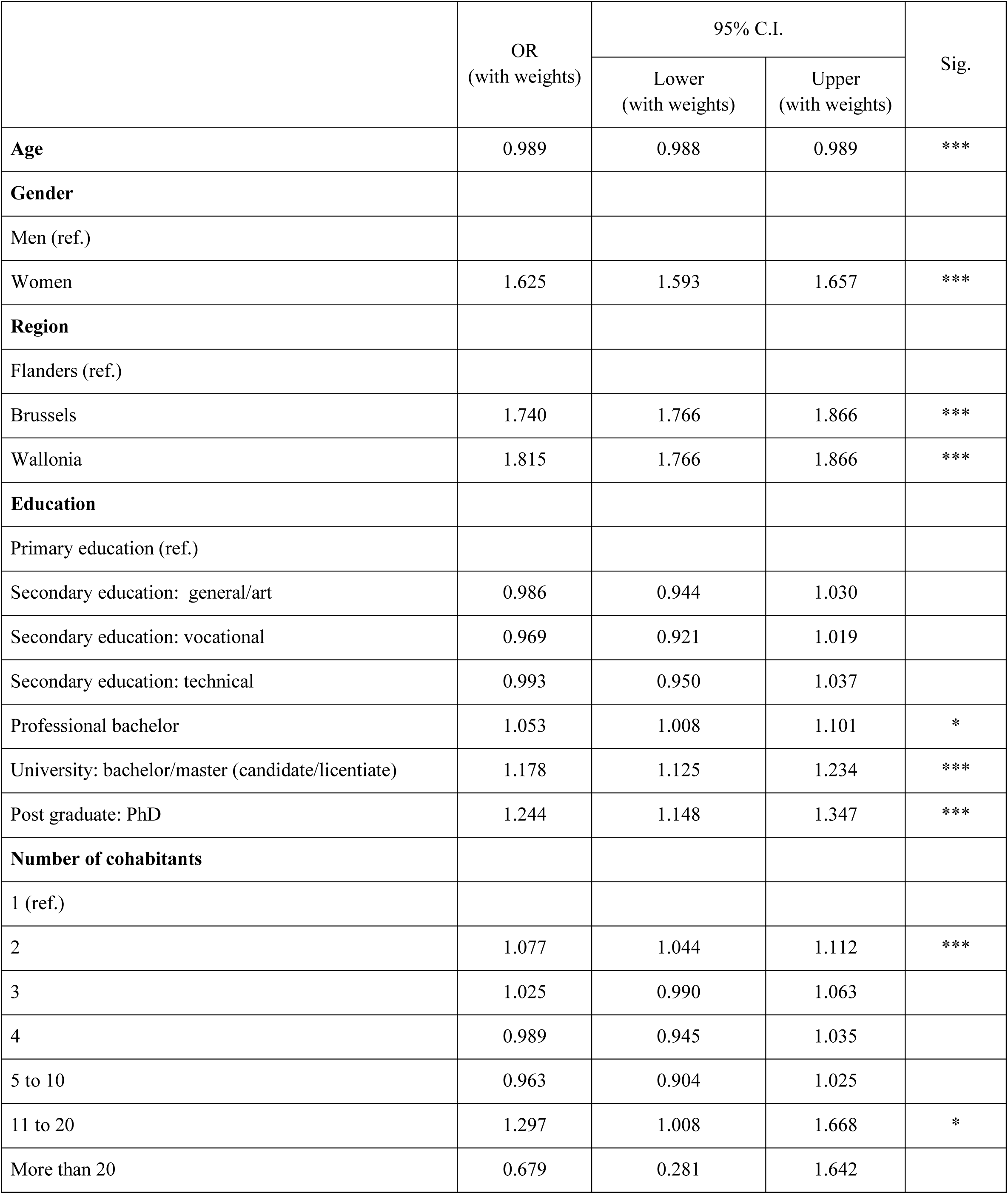
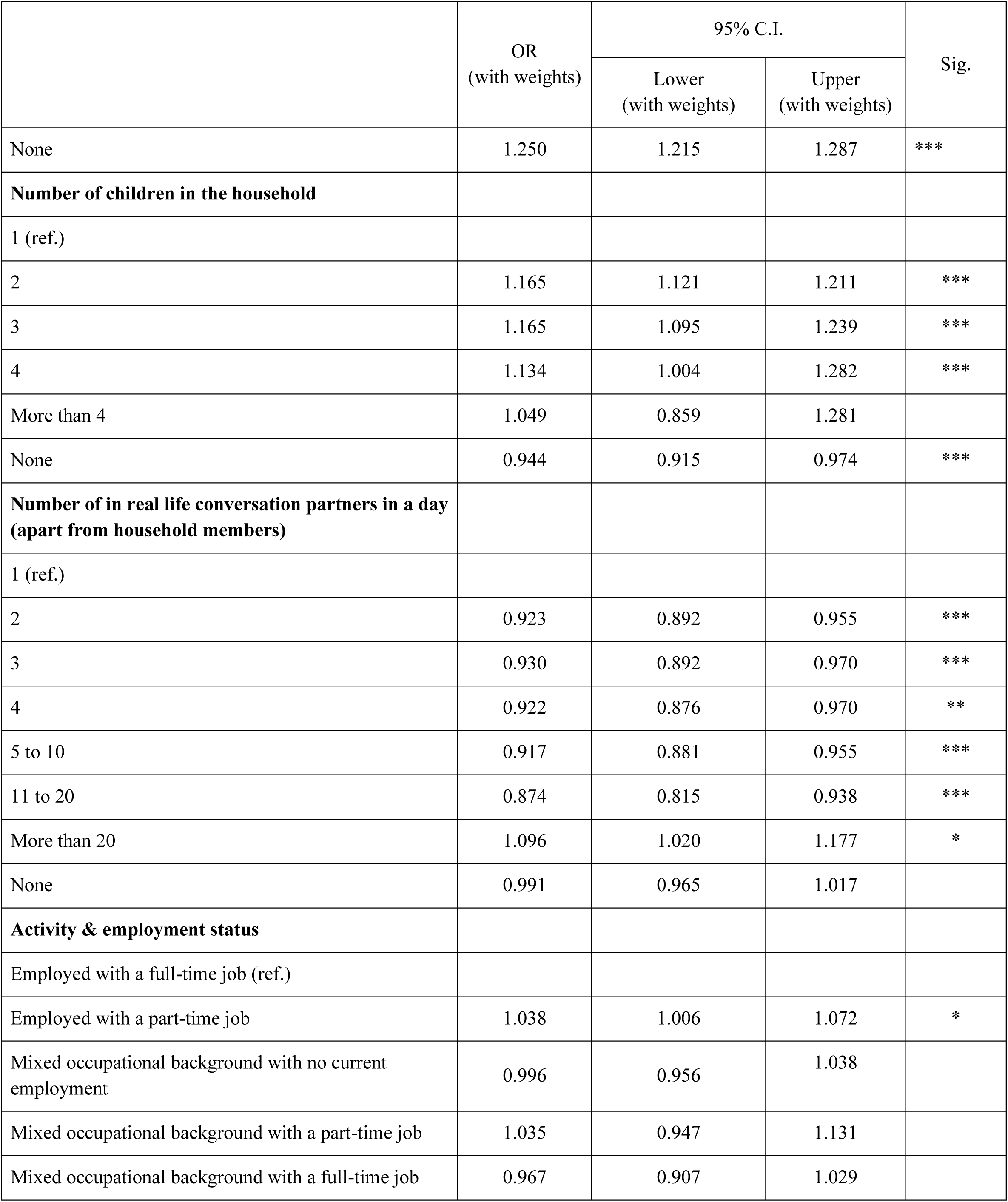
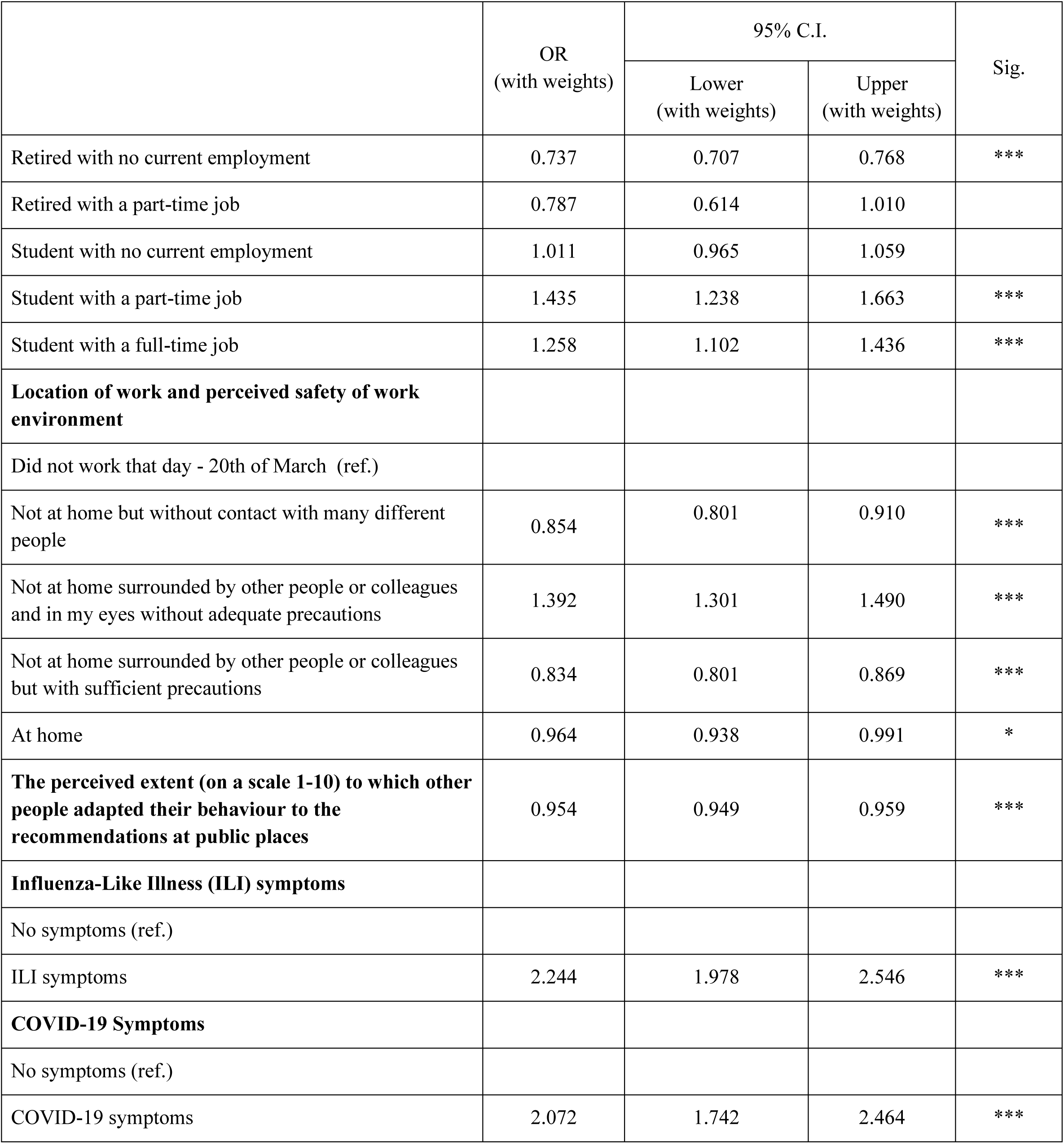
Results of the generalized linear model applying survey weights.

|  | OR<br>(with weights) | 95% C.I. |  | Sig. |
| --- | --- | --- | --- | --- |
|  |  | Lower<br>(with weights) | Upper<br>(with weights) |  |
| **Age** | 0.989 | 0.988 | 0.989 | *** |
| **Gender** |  |  |  |  |
| Men (ref.) |  |  |  |  |
| Women | 1.625 | 1.593 | 1.657 | *** |
| **Region** |  |  |  |  |
| Flanders (ref.) |  |  |  |  |
| Brussels | 1.740 | 1.766 | 1.866 | *** |
| Wallonia | 1.815 | 1.766 | 1.866 | *** |
| **Education** |  |  |  |  |
| Primary education (ref.) |  |  |  |  |
| Secondary education: general/art | 0.986 | 0.944 | 1.030 |  |
| Secondary education: vocational | 0.969 | 0.921 | 1.019 |  |
| Secondary education: technical | 0.993 | 0.950 | 1.037 |  |
| Professional bachelor | 1.053 | 1.008 | 1.101 | * |
| University: bachelor/master (candidate/licentiate) | 1.178 | 1.125 | 1.234 | *** |
| Post graduate: PhD | 1.244 | 1.148 | 1.347 | *** |
| **Number of cohabitants** |  |  |  |  |
| 1 (ref.) |  |  |  |  |
| 2 | 1.077 | 1.044 | 1.112 | *** |
| 3 | 1.025 | 0.990 | 1.063 |  |
| 4 | 0.989 | 0.945 | 1.035 |  |
| 5 to 10 | 0.963 | 0.904 | 1.025 |  |
| 11 to 20 | 1.297 | 1.008 | 1.668 | * |
| More than 20 | 0.679 | 0.281 | 1.642 |  |
|  |  | Lower<br>(with weights) | Upper<br>(with weights) |  |
| None | 1.250 | 1.215 | 1.287 | *** |
| **Number of children in the household** |  |  |  |  |
| 1 (ref.) |  |  |  |  |
| 2 | 1.165 | 1.121 | 1.211 | *** |
| 3 | 1.165 | 1.095 | 1.239 | *** |
| 4 | 1.134 | 1.004 | 1.282 | *** |
| More than 4 | 1.049 | 0.859 | 1.281 |  |
| None | 0.944 | 0.915 | 0.974 | *** |
| **Number of in real life conversation partners in a day**<br>**(apart from household members)** |  |  |  |  |
| 1 (ref.) |  |  |  |  |
| 2 | 0.923 | 0.892 | 0.955 | *** |
| 3 | 0.930 | 0.892 | 0.970 | *** |
| 4 | 0.922 | 0.876 | 0.970 | ** |
| 5 to 10 | 0.917 | 0.881 | 0.955 | *** |
| 11 to 20 | 0.874 | 0.815 | 0.938 | *** |
| More than 20 | 1.096 | 1.020 | 1.177 | * |
| None | 0.991 | 0.965 | 1.017 |  |
| **Activity & employment status** |  |  |  |  |
| Employed with a full-time job (ref.) |  |  |  |  |
| Employed with a part-time job | 1.038 | 1.006 | 1.072 | * |
| Mixed occupational background with no current employment | 0.996 | 0.956 | 1.038 |  |
| Mixed occupational background with a part-time job | 1.035 | 0.947 | 1.131 |  |
| Mixed occupational background with a full-time job | 0.967 | 0.907 | 1.029 |  |
|  |  | Lower<br>(with weights) | Upper<br>(with weights) |  |
| Retired with no current employment | 0.737 | 0.707 | 0.768 | *** |
| Retired with a part-time job | 0.787 | 0.614 | 1.010 |  |
| Student with no current employment | 1.011 | 0.965 | 1.059 |  |
| Student with a part-time job | 1.435 | 1.238 | 1.663 | *** |
| Student with a full-time job | 1.258 | 1.102 | 1.436 | *** |
| **Location of work and perceived safety of work environment** |  |  |  |  |
| Did not work that day - 20th of March (ref.) |  |  |  |  |
| Not at home but without contact with many different people | 0.854 | 0.801 | 0.910 | *** |
| Not at home surrounded by other people or colleagues and in my eyes without adequate precautions | 1.392 | 1.301 | 1.490 | *** |
| Not at home surrounded by other people or colleagues but with sufficient precautions | 0.834 | 0.801 | 0.869 | *** |
| At home | 0.964 | 0.938 | 0.991 | * |
| **The perceived extent (on a scale 1-10) to which other people adapted their behaviour to the recommendations at public places** | 0.954 | 0.949 | 0.959 | *** |
| **Influenza-Like Illness (ILI) symptoms** |  |  |  |  |
| No symptoms (ref.) |  |  |  |  |
| ILI symptoms | 2.244 | 1.978 | 2.546 | *** |
| **COVID-19 Symptoms** |  |  |  |  |
| No symptoms (ref.) |  |  |  |  |
| COVID-19 symptoms | 2.072 | 1.742 | 2.464 | *** |

## Discussion

This study examined potential predictors of psychological distress during the initial COVID-19 lockdown in Belgium, leveraging data from the Great Corona Study (GCS). With its large-scale, real-time data collection, the GCS provided a unique opportunity to explore the early mental health impacts of the pandemic. Some of these observations were communicated in broad terms in the Belgian press the day after data collection, but had hitherto not been analyzed in depth. Using a combination of random forest analysis and literature review, we identified key sociodemographic, occupational, and behavioral factors associated with psychological distress, measured via the GHQ-12 questionnaire. Our findings highlight several significant associations. Younger individuals and women were at a higher risk of experiencing distress, as were those living in Brussels and Wallonia. Household composition played a notable role, with individuals living with two, or with eleven to twenty cohabitants or multiple children showing increased odds of distress. Those with frequent real-life interactions exhibited lower distress levels. Occupational factors were also influential, with part-time employees and working students facing higher distress levels. Conversely, retired individuals with no part-time employment and those perceiving their work environments as safe had reduced odds of psychological distress. Additionally, compliance with public health measures and perceptions of others’ adherence were linked to mental well-being. Finally, experiencing Influenza-Like Illness (ILI) or COVID-19 symptoms was strongly associated with elevated distress.

The results of this study align with the broader body of evidence presented in the introduction, most of which relates to data collected in later stages of the pandemic or focused on specific subgroups of the population. Our results support the well-documented sociodemographic disparities in mental health outcomes. Women and young adults emerged as one of the most vulnerable groups, similarly to findings from previously published research highlighting their heightened levels of psychological distress during the early stages of the pandemic (7, 9). The role of social determinants, such as educational level and household composition also are in alignment with the current literature (3). Our analysis further emphasizes the connection between in-person interactions and mental well-being, reinforcing previously published research that considered social isolation to be a key driver of psychological distress, especially among young people (9). At this early stage of the pandemic, it was indeterminable by our analyses whether fewer social contacts caused higher distress levels, or vice versa.

In addition to confirming existing knowledge on psychological distress during the early phase of the COVID-19 pandemic, our results provide new insights into key factors that shaped mental health outcomes. One of the most striking findings was the exceptionally high odds ratios (ORs) for experiencing influenza-like illness (ILI) and COVID-19 symptoms. These high ORs underscore the significant health risks present at the time and highlight the extent to which physical health concerns may have contributed to psychological distress. At the same time, an also influential factor was how people perceived others’ adaptation to COVID-19 measures, both in public spaces and in the workplace. Our findings show that individuals who observed stronger adherence to preventive measures reported lower levels of distress. This suggests that beyond personal behavior, the social environment and regional differences also played a crucial role in shaping psychological well-being during the crisis. Similarly, work location was an important factor, as those working on-site faced different psychological stressors compared to those working remotely. The extent to which coworkers followed preventive measures also influenced mental health, emphasizing the interconnected nature of compliance, risk perception, and well-being.

A key strength of this study is the use of data from the Great Corona Study (GCS). The GCS offers several advantages, including its exceptionally large sample size, which enhances the statistical power and generalizability of our findings. Furthermore, while previous research has extensively examined epidemiological trends, economic consequences, and behavioral adaptations during the pandemic (18–20), the psychological dimension remains relatively underexplored. By focusing on mental health outcomes, particularly in relation to perceived adherence to COVID-19 measures and work location, this study fills an important gap in the literature. The findings highlight how individuals’ psychological well-being was shaped not only by personal experiences but also by their perceptions of others’ behaviors, offering novel insights into the social and environmental determinants of distress.

Additionally, the study benefits from its ability to assess both individual and contextual factors influencing mental health. The strong associations observed between ILI and COVID-19 symptoms with psychological distress emphasize the role of health-related uncertainty, while the impact of perceived public and workplace adherence to preventive measures underscores the significance of collective behaviors. These insights provide valuable evidence for policymakers, emphasizing the importance of fostering visible and consistent public health compliance to mitigate psychological distress during future crises.

Our study successfully explored a uniquely broad selection of predictor variables in the early stages of the COVID-19 pandemic. However, like any study, ours also has certain limitations.

First, our study sample is not fully representative of the entire Belgian population. For instance, most respondents were from Flanders, leading to an underrepresentation of individuals from Wallonia and Brussels. Additionally, the gender distribution was not balanced, as female respondents outnumbered male participants. Age group representation is also imbalanced, particularly among children (below the age of 18), whose responses were collected and recorded by their parents, resulting in a relatively small subgroup (1.3% of the participants). Furthermore, there are deviations in educational level distribution compared to the overall Belgian population. To address these discrepancies, we applied survey weights in our analysis. However, future research could benefit from conducting similar analyses on smaller, more specific subgroups of the Great Corona Study respondents. At the same time, it is important to acknowledge that the GCS is an observational study based on voluntary questionnaire participation, which introduces an inherent bias that cannot be fully corrected using post-stratification weights alone.

An additional consideration pertains to the GHQ-12 cutoffs. Thresholds in research and clinical practice commonly range from 1/2 to 6/7, with the 2/3 cutoff being the most frequently applied (21), particularly in research settings (23–25). The literature however does not establish a definitive threshold distinguishing the 2/3 and 3/4 divisions in research, as evidence supports both approaches (24–27). In this study, we selected the cutoff that we deemed most appropriate based on existing research (23–25, 28). However, we acknowledge that the alternative cutoff method is also valid. To assess the potential impact of this choice, we conducted the analysis using both cutoff criteria, which yielded no differences in the overall conclusions.

Another major limitation of this study is the lack of data on preexisting mental health conditions among respondents. This is particularly important, as evidence suggests that individuals with prior mental health conditions were especially vulnerable to heightened psychological distress during the pandemic (3, 6). Additionally, in this GCS wave, we do not have information on respondents’ levels of social support or perceived loneliness, both of which are known to be significant contributors to mental well-being in crisis situations (3, 4, 8, 9).

Finally, a limitation of our analysis is that we did not account for potential interaction effects between predictor variables. While our approach focused on identifying the most relevant individual predictors using random forest analysis and subsequently selecting variables for the generalized linear model, interactions between variables were not explicitly modeled. Future research could explore whether incorporating interaction terms enhances the explanatory power and accuracy of the model.

## Conclusions

This study provides valuable insights into the psychological distress experienced during the first week of the COVID-19 lockdown in Belgium. By leveraging data from the Great Corona Study (GCS), we identified key sociodemographic, occupational, and health-related factors associated with mental health outcomes. Our findings confirm existing evidence on vulnerable groups, particularly young adults and women, while also highlighting the role of household composition, work environment, and perceived adherence to public health measures in influencing psychological distress.

Beyond affirming known predictors, our study underlines the significant association between experiencing influenza-like illness or COVID-19 symptoms and heightened psychological distress, emphasizing the interaction between physical and mental health during a public health crisis. Additionally, the perception of others’ compliance with preventive measures emerged as a critical factor, suggesting that risk perception strongly influenced psychological well-being at this early stage of the pandemic.

While the study benefits from a large, diverse dataset and a robust analytical approach, limitations such as sampling biases, the absence of preexisting mental health data, and unaccounted interaction effects should be considered. Despite these constraints, our findings contribute to a more comprehensive understanding of the mental health impact of pandemic-related stressors. Future research should further explore the interplay between and evolution of social, occupational, and health-related factors. This can then be put to use to inform the design of interventions, in order to limit the mental well-being impact during future public health emergencies.

## Ethics approval

The Great Corona Study was reviewed and approved by the Ethics Committee of the University Hospital Antwerp – University of Antwerp (Commissie voor Medische Ethiek UZA– UAntwerpen; approval number EC 20/13/146).

## Consent for publication

Not applicable.

## Availability of data and materials

Data are not publicly available; data from the Great Corona Study can be requested via email (https://www.uantwerpen.be/en/projects/great-corona-study/), and the Great Corona Study team will consider data sharing.

## Author’s contributions

NH, PB, and KP conceived and designed the study and collected the data. ZZ performed the statistical analysis and interpreted the results under the supervision of NH. ZZ drafted the first version of the manuscript. NH, KVDB, PB, and KP critically reviewed the manuscript and provided important intellectual revisions. All authors read and approved the final manuscript.

## Acknowledgements

P.B and N.H. acknowledge funding from the Methusalem-Centre of Excellence consortium VAX-IDEA and the University of Antwerp Research Fund.

Z.Z. acknowledges support from a Methusalem research grant from the Flemish government. The authors acknowledge Jonas Crèvecoeur for maintaining the Great Corona Study database, Yessica Adelwin Natalia for her help with data access and handling, and for dr. Lisa Hermans for her help with constructing survey weights.

## Appendix

## Appendix A Variable Importance Plots corresponding to the Random Forest Analysis conducted with 1.000 and 10.000 random observations

**Figure 1:**
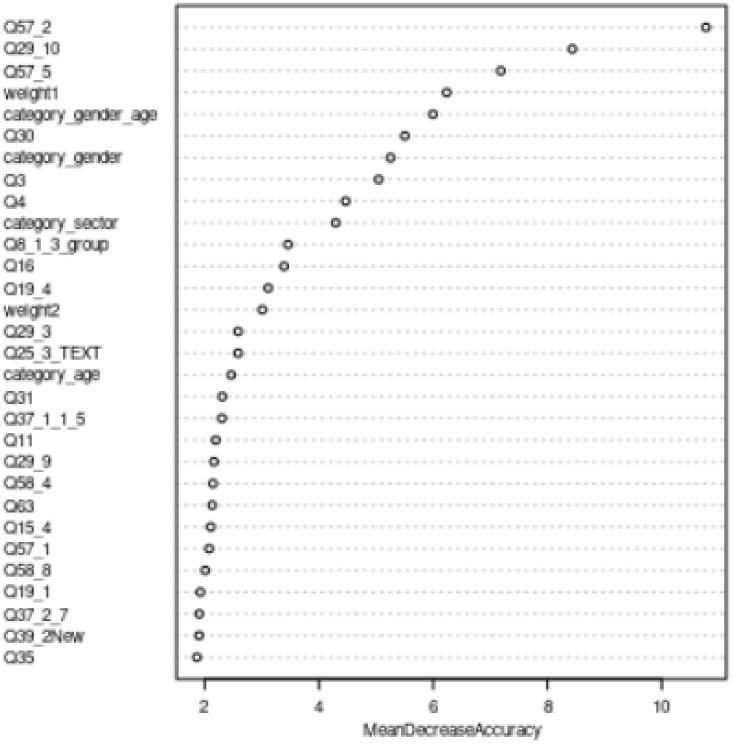
Mean Decrease Accuracy with 1.000 random observations

**Figure 2:**
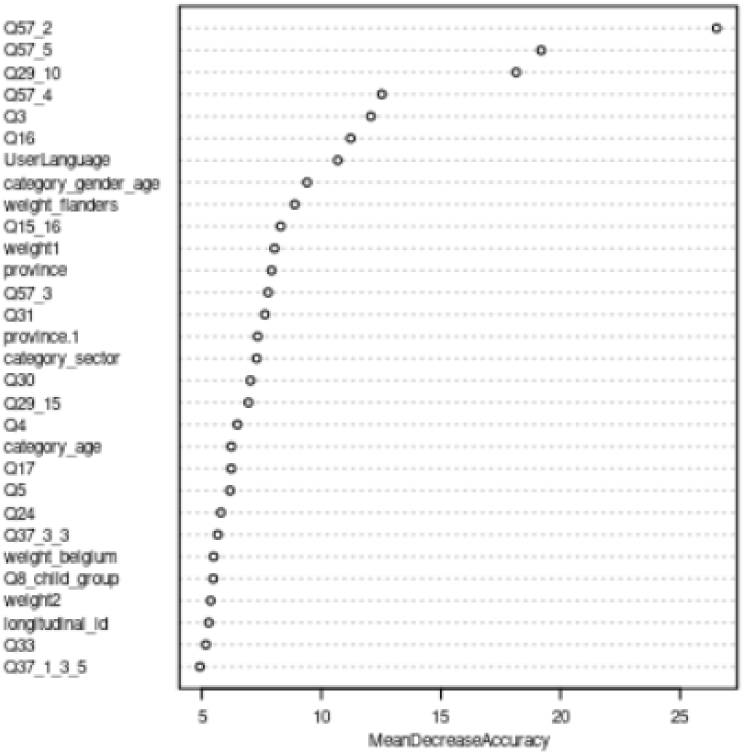
Mean Decrease Accuracy with 10.000 random observations

**Figure 3:**
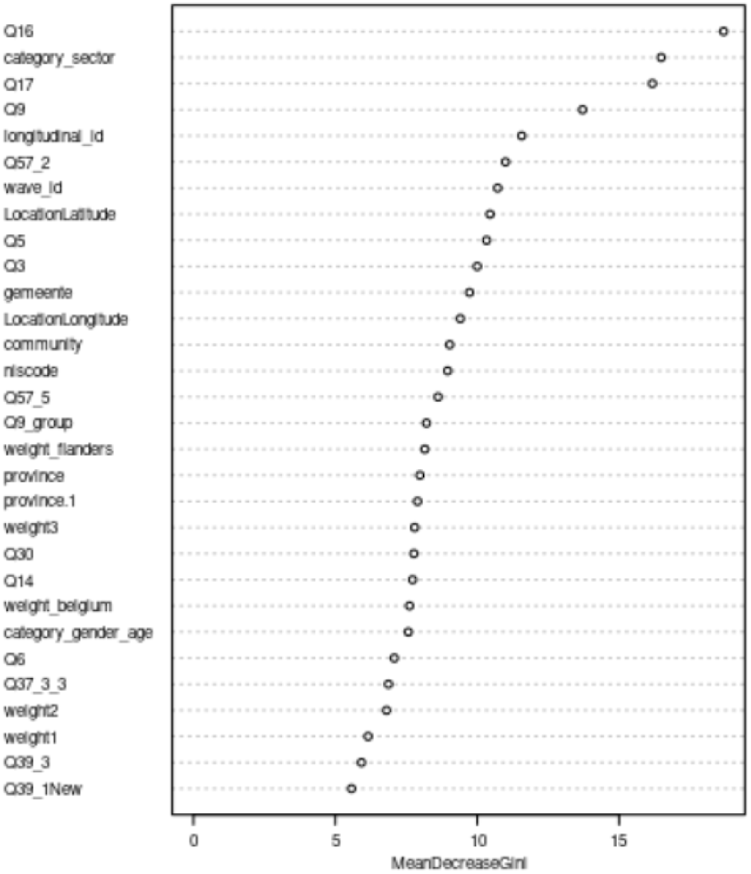
Mean Decrease Gini with 1.000 random observations

**Figure 4:**
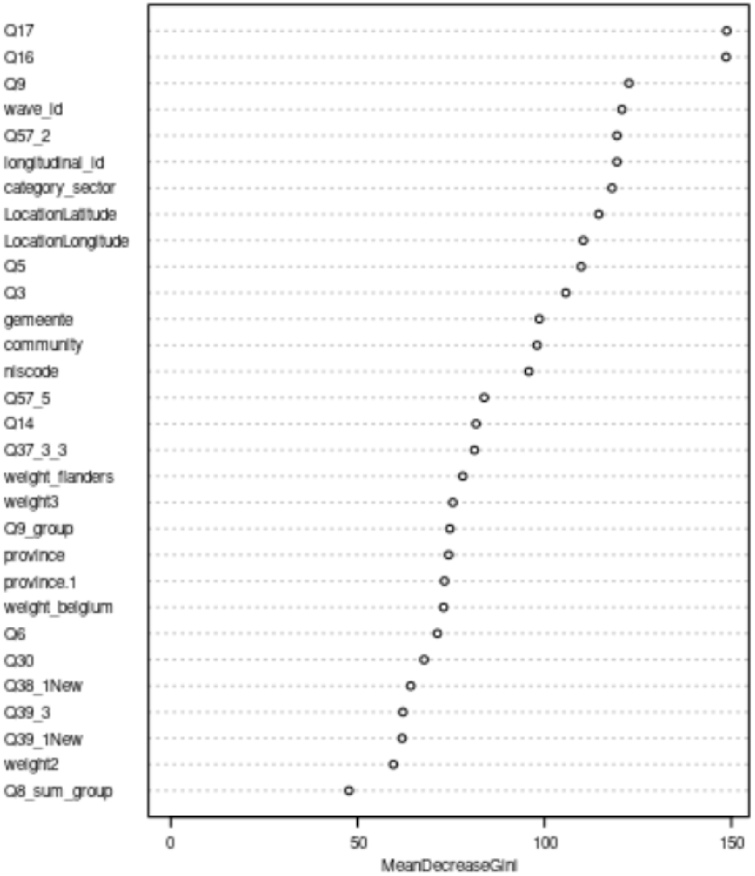
Mean Decrease Gini with 10.000 random observations

## Appendix B Characteristics of the study sample

**Table 1-.** Characteristics of the study sample.

| Variable | Level | GHQ-12 Low (n, %) | GHQ-12 High (n, %) | Total (n, %) |
| --- | --- | --- | --- | --- |
| **Age** | 0-17 year | 1,856<br>(42.63%) | 2,498<br>(57.37%) | 4,354<br>(1.31%) |
|  | 18-35 year | 56,359<br>(39.98%) | 84,618<br>(60.02%) | 140,977<br>(42.44%) |
|  | 36-65 year | 82,635<br>(48.20%) | 88,810<br>(51.80%) | 171,445<br>(51.61%) |
|  | 66+ year | 9,967<br>(64.75%) | 5,426<br>(35.25%) | 15,393<br>(4.63%) |
| **Gender** | Men | 61,539<br>(53.84%) | 52,751<br>(46.16%) | 114,290<br>(34.41%) |
|  | Women | 89,278<br>(40.98%) | 128,601<br>(59.02%) | 217,879<br>(65.59%) |
| **Region** | Flanders | 140,950<br>(46.83%) | 160,043<br>(53.17%) | 300,993<br>(90.61%) |
|  | Brussels | 4,255<br>(31.46%) | 9,270<br>(68.54%) | 13,525<br>(4.07%) |
|  | Wallonia | 5,612<br>(31.79%) | 12,039<br>(68.21%) | 17,651<br>(5.31%) |
| **Education** | Primary education (ref.) | 2,116<br>(51.65%) | 1,981<br>(48.35%) | 4,097<br>(1.23%) |
|  | Secondary education:<br>general/art | 14,999<br>(49.75%) | 15,149<br>(50.25%) | 30,148<br>(9.08%) |
|  | Secondary education:<br>vocational | 8,327<br>(48.66%) | 8,785<br>(51.34%) | 17,112<br>(5.15%) |
|  | Secondary education:<br>technical | 19,258<br>(51.09%) | 18,436<br>(48.91%) | 37,694<br>(11.35%) |
|  | Professional bachelor | 52,808<br>(45.98%) | 62,054<br>(54.02%) | 114,862<br>(34.58%) |
|  | University:<br>bachelor/master<br>(candidate/licentiate) | 49,549<br>(41.62%) | 69,498<br>(58.38%) | 119,047<br>(35.84%) |
|  | Post graduate: PhD | 3,760<br>(40.83%) | 5,449<br>(59.17%) | 9,209<br>(2.77%) |
| **Number of cohabitants** | 1 (ref.) | 50,272<br>(50.60%) | 49,083<br>(49.40%) | 99,355<br>(29.91%) |
|  | 2 | 27,718<br>(43.86%) | 35,474<br>(56.14%) | 63,192<br>(19.02%) |
|  | 3 | 34,920<br>(42.76%) | 46,745<br>(57.24%) | 81,665<br>(24.59%) |
|  | 4 | 14,316 | 19,070 | 33,386 |
|  |  | (42.88%) | (57.12%) | (10.05%) |
|  | 5 to 10 | 5,584<br>(41.98%) | 7,717<br>(58.02%) | 13,301<br>(4.00%) |
|  | 11 to 20 | 238<br>(38.70%) | 377<br>(61.30%) | 615<br>(0.19%) |
|  | More than 20 | 11<br>(45.83%) | 13<br>(54.17%) | 24<br>(0.01%) |
|  | None | 17,758<br>(43.71%) | 22,873<br>(56.29%) | 40,631<br>(12.23%) |
| **Number of children in the household** | 1 (ref.) | 21,702<br>(42.82%) | 28,979<br>(57.18%) | 50,681<br>(15.26%) |
|  | 2 | 24,883<br>(41.69%) | 34,800<br>(58.31%) | 59,683<br>(17.97%) |
|  | 3 | 6,475<br>(41.79%) | 9,019<br>(58.21%) | 15,494<br>(4.66%) |
|  | 4 | 1,070<br>(41.93%) | 1,482<br>(58.07%) | 2,552<br>(0.77%) |
|  | More than 4 | 398<br>(42.48%) | 539<br>(57.52%) | 937<br>(0.28%) |
|  | None | 96,289<br>(47.47%) | 106,533<br>(52.52%) | 202,822<br>(61%) |
| **Number of in real life conversation partners in a day (apart from household members)** | 1 (ref.) | 27,611<br>(45.47%) | 33,110<br>(54.52%) | 60,721<br>(18.28%) |
|  | 2 | 20,175<br>(47.73%) | 22,086<br>(52.26%) | 42,261<br>(12.72%) |
|  | 3 | 10,653<br>(47.98%) | 11,547<br>(52.01%) | 22,200<br>(6.68%) |
|  | 4 | 6,446<br>(48%) | 6,983<br>(51.99%) | 13,429<br>(4.04%) |
|  | 5 to 10 | 14,438 | 15,115 | 29,553<br>(8.89%) |
|  | 11 to 20 | 3,769<br>(48.24%) | 4,043<br>(51.73%) | 7,812<br>(2.35%) |
|  | More than 20 | 3,210<br>(43%) | 4,254<br>(51.75%) | 7,464<br>(2.24%) |
|  | None | 64,515 | 84,214 | 148,729<br>(44.77%) |
| **Activity and employment status** | Employed with a full-time job (ref.) | 86,618<br>(45.73%) | 102,770<br>(54.26%) | 189,388<br>(57%) |
|  | Employed with a part-time job | 21,537<br>(42.46%) | 29,182<br>(57.53%) | 50,719<br>(15.26%) |
|  | Mixed occupational background with no current employment | 8,519<br>(45.5%) | 10,200<br>(54.49%) | 18,719<br>(5.63%) |
|  | Mixed occupational background with a part-time job | 1,595<br>(42.74%) | 2,136<br>(57.25%) | 3,731<br>1.12% |
|  | Mixed occupational background with a full-time job | 3,588<br>(46.15%) | 4,185<br>(53.84%) | 7,773<br>(2.34%) |
|  | Retired with no current employment | 16,319<br>(62.71%) | 9,703<br>(37.28%) | 26,022<br>(7.83%) |
|  | Retired with a part-time job | 163<br>(59.27%) | 112<br>(40.72%) | 275<br>(0.08%) |
|  | Student with no current employment | 10,972<br>(35.45%) | 19,974<br>(64.54%) | 30,946<br>(9.31%) |
|  | Student with a part-time job | 616<br>(32.25%) | 1,294<br>(67.74%) | 1,910<br>(0.57%) |
|  | Student with a full-time job | 890<br>(33.13%) | 1,796<br>(66.86%) | 2,686<br>(0.80%) |
| **Location of work and perceived safety of work environment** | Did not work that day - 20th of March (ref.) | 74,934<br>(45.34%) | 90,333<br>(54.66%) | 165,267<br>(49.75%) |
|  | Not at home but without contact with many different people | 4,562<br>(52.30%) | 4,161<br>(47.70%) | 8,723<br>(2.63%) |
|  | Not at home surrounded | 2,748 | 4,670 | 7,418 |
|  | by other people or colleagues and in my eyes without adequate precautions | (37.05%) | (62.95%) | (2.23%) |
|  | Not at home surrounded by other people or colleagues but with sufficient precautions | 15,183<br>(50.03%) | 15,163<br>(49.97%) | 30,346<br>(9.14%) |
|  | At home | 53,390<br>(44.34%) | 67,025<br>(55.66%) | 120,415<br>(36.25%) |
| **The perceived extent (on a scale 1-10) to which other people adapted their behaviour to the recommendations at public places** | 1-5 | 10,324<br>(40.11%) | 15,415<br>(59.89%) | 25,739<br>(0.55%) |
|  | 6-10 | 140,493<br>(45.85%) | 165,937<br>(54.15%) | 306,430<br>(99.45%) |
| **Influenza-Like Illness (ILI) symptoms** | No symptoms (ref.) | 150,124<br>(45.63%) | 178,881<br>(54.37%) | 329,005<br>(99.05%) |
|  | ILI symptoms | 693<br>(21.90%) | 2,471<br>(78.10%) | 3,164<br>(0.95%) |
| **COVID-19 Symptoms** | No symptoms (ref.) | 150,463<br>(45.55%) | 179,885<br>(54.45%) | 330,348<br>(99.45%) |
|  | COVID-19 symptoms | 354<br>(19.44%) | 1,467<br>(80.56%) | 1,821<br>(0.55%) |

